# Clinical outcomes of 402 patients with COVID-2019 from a single center in Wuhan, China

**DOI:** 10.1101/2020.03.07.20032672

**Authors:** Yingjie Wu, Wei Guo, Huan Liu, Bei Qi, Ke Liang, Haibo Xu, Zhiyong Peng, Shu-Yuan Xiao

## Abstract

The SARS-CoV-2 outbreak is causing widespread infections and significant mortality. Previous studies describing clinical characteristics of the disease contained small cohorts from individual centers or larger series consisting of mixed cases from different hospitals. We report analyses of mortality and disease severity among 402 patients from a single hospital. The cohort included 297 patients with confirmed and 105 with suspected diagnosis. The latter group met the criteria for clinical diagnosis but nucleic acid tests results were initially interpreted as suspicious. Data were compared between genders and among different age groups. The overall case fatality is 5.2%. However, patients 70 years of age or older suffered a significantly higher mortality (17.8%), associated with more patients having severe or critical illness (57.5%). Patients 50 years of age or older had a mortality of 8.0%, and those younger than 50 years, 1.2%. Male patients had a mortality of 7.6% versus 2.9% in females.

## Introduction

In December 2019, a cluster of “atypical” pneumonia cases with then unknown causes occurred in several hospitals in Wuhan, Hubei Province, China[2]. Most of the initial patients had fever, fatigue and non-productive cough, and showed a characteristic ground glass shadow on chest CT imaging of the lungs. Some of these patients could be linked to a local fresh seafood market, Huanan Seafood Market, although others could not. A coronavirus was subsequently isolated and genomically sequenced. It was found that the viruses share nucleotide sequence homology of 79.5% to SARS-CoV and 85-96% to bat SARS-like coronavirus bat-SL-CoVZC45 at the whole genome level[3]. The virus was initially named 2019-novel coronavirus(2019-nCoV) on January 12, and subsequently, SARS-CoV-2 on February 11. Disease caused by the infection is now designated coronavirus infected disease 2019, or COVID-19. The outbreak thus represents a new emerging viral disease due to species “jumping” of an animal virus to humans. Currently, human-to-human transmissions of the virus have reached an unprecedent magnitude, in community, healthcare facilities, and at homes[4], and spread to entire China and some parts of the world.

Initially, recognition and diagnosis of the disease, namely COVID-19, were based on the characteristic clinical, laboratory and radiological findings, with exclusion of other known respiratory agents. Soon after, definitive diagnosis required the detection of viral sequence by a nucleic acid test, reverse-transcriptase polymerase chain reaction (RT-PCR). Most previous reports on clinical case studies were based on this definition. However, it became evident that a significant portion of cases showed negative viral detections in pharyngeal swab specimens, although tested repeatedly, but clinically fit the diagnosis. The possible causes of this discrepancy are several, including but not limited to (1) not all patients with the lower respiratory tract involvement shed virus from the upper respiratory tracts, at least early on; (2) there might be insufficient consistency in sampling; (3) the sensitivity and specificity of the nucleic acid tests had not been sufficiently investigated[5]. Strictly following this criterium had prevented many patients from receiving timely care, particularly early on when availability of enough test kits could not meet the demand of the large number of symptomatic patients. The clinicians and authorities recognized these problems and made prompt changes to the diagnostic guidelines, so that patients meeting the criteria for clinical diagnosis in Hubei Province were treated as COVID-19 patients. In the updated version of the guidelines for clinical diagnosis and management of COVID-19 by the National Health Commission of China, definition for clinical diagnosis does not require a nucleic acid test result.

There have been several studies describing the clinical characteristics of SARS-CoV-2 infected patients[1, 6, 7], including symptoms, lab tests and radiographic features. These were smaller series, from 41[6] to 138 confirmed cases[1]. Some of the larger series consisted of mixed cases submitted from hospitals of varying sizes and settings[7]. Analysis of larger series with cases from a single center and expanding a longer period of time should offer more accurate information about the overall clinical outcomes, mortality and morbidity, because these cases would have followed more or less uniform diagnostic algorithms and had received more consistent treatment regimens. The results should have less interference from uncontrollable factors such as inconsistency in reporting from individual hospitals.In addition, patients included in these prior studies all had diagnosis confirmed by the nucleic acid tests for pharyngeal swab specimens. For the reasons described above, some patients not included due to suspicious nucleic acid test results may represent more mild illness, thus causing bias in clinical outcome analysis. Therefore, it is important to include cases with typical clinical presentations and course, even though “suspicious” result on nucleic acid tests, in studies of clinical outcomes and disease characteristics. In the current study, we analyzed data on 402 patients from a single hospital from December 2019 to February 2, 2020, with emphasis on mortality in these patients, in hope to understand characteristics related to clinical outcome in a more uniform clinical setting.

## Material and Methods

### Patients

Patients presented to our hospital and had nucleic acid tests showing “positive” or “suspicious” results from December 2019 to February 2, 2020 were included in this study. Electronic medical charts were reviewed. Patients demographics, status of nucleic acid tests, time of presentation and or illness, length of symptoms, and so on, were recorded. The clinical severity status (ie, common/mild, severe, or critical) and death were monitored up to February 2, 2020, the final date of follow-up.

All patients met the criteria for clinical diagnosis given by The National Health Commission of China (NHCC) Guidelines on novel coronavirus pneumonia for diagnosis and disease severity triage (5^th^ Edition). Briefly, diagnosis was based on epidemic exposure, plus two of the following clinical findings: fever, radiographic features, normal or lowered white blood cells (WBC) or reduced lymphocyte count.

### Interpretation of results for real-time Reverse Transcription Polymerase Chain Reaction Assay for SARS-CoV-2

The real-time Reverse Transcription Polymerase Chain Reaction using the laryngeal swabs were performed as reported previously[1]. Initially, results of “positive” nucleic acid tests were defined as 2 amplification sites in quantitative RT-PCR, while the “suspicious” results were defined as one of the two sites had a positive signal. Along with accumulating experience and knowledges about this disease and the test, both scenarios were considered as “positive” subsequently. Therefore, all cases in this study that had been classified as “suspicious” were in fact positive cases. In addition, they all met the criterial for clinical diagnosis. In the following analysis, we keep the label of “suspicious” but also analyzed the initially confirmed cases in a separate group, side-by-side.

### Severity Group Designation

The common (mild) cases were those only had fever, respiratory symptoms, and pneumonia on chest radiography. Severe cases need to meet one of the following criteria: (1) respiratory distress, RR>=30/min; (2) resting blood oxygen saturation =< 93%; or (3) arterial blood oxygen partial pressure (PaO2)/FiO2 =<300 mmHg. Critical cases meet one of the following: (1) respiratory failure needing mechanical oxygenation; (2) shock; or (3) development of other organ failure, requiring intensive care unit (ICU) care. Around 70-80% of patients were mild, and 20-30% were severe or critical.

### Statistical Analysis

Categorical variables were described as frequency rates and percentages. Proportions for categorical variables were compared using the χ^2^ test. All statistical analyses were performed using GraphPad Prism (GraphPad Company, San Diego, CA, USA) version 6.0 software. P-value “< 0.05” was considered with statistical significance.

This study was approved by the Ethics Board of Zhongnan Hospital of Wuhan University (No.2020012).

## Results

### Case fatality analysis of SARS-CoV-2 infected patients

As showed in Table 1 and Figure 1A, the fatality of all confirmed and suspected COVID-19 patients was 5.2%, while fatality of confirmed cases was 5.7%. Male patients had higher fatality (7.6%) than females (2.9%) among all patients (P=0.04) (Figure 1B), while in confirmed cases, fatality in males was 8.8% and female 2.7% (P=0.03) (Figure 1C). The classification of case fatalities among different age groups were shown in Table 1 and Figure 1D and E. Fatalities in patients younger than 30 years of age, 30 to 49, 50 to 69, 70 and older were 0, 1.5%, 3.6% and 17.8% respectively among all suspected and confirmed patients (Figure 1D). The fatalities of patients under 30, from 30 to 49, from 49 to 69, 70 and older, were 0, 1.0%, 4.2% and 20.0%, respectively, among confirmed patients (Figure 1E). Taking 50 years old as a cutoff in confirmed cases, the mortality in patients over 50 years old was 14.3% in males and 4.5% in females (**Table 1**).

**Table 1.** Case fatalities by age and gender among patients with suspected and confirmed diagnosis for SARS-CoV-2 infection.

| Gender and Age<br>groups | Confirmed and suspected |  |  | Confirmed |  |  |
| --- | --- | --- | --- | --- | --- | --- |
|  | No. of patients | Death | Fatality (%) | No. of patients | Death | Fatality (%) |
| Gender |  |  |  |  |  |  |
| Male | 198 | 15 | 7.6 | 147 | 13 | 8.8 |
| Female | 204 | 6 | 2.9 | 150 | 4 | 2.7 |
| Total | 402 | 21 | 5.2 | 297 | 17 | 5.7 |
| Age ($\geq 50$ years) | | | | | | |
| Male | 115 | 14 | 12.2 | 84 | 12 | 14.3 |
| Female | 123 | 5 | 4.1 | 89 | 4 | 4.5 |
| Age (years) |  |  |  |  |  |  |
| $\leq 2$ | 2 | 0 | 0.0 | 2 | 0 | 0.0 |
| $> 2 < 30$ | 29 | 0 | 0.0 | 18 | 0 | 0.0 |
| $\geq 30 < 50$ | 133 | 2 | 1.5 | 104 | 1 | 1.0 |
| $\geq 50 < 70$ | 165 | 6 | 3.6 | 118 | 5 | 4.2 |
| $\geq 70$ | 73 | 13 | 17.8 | 55 | 11 | 20.0 |

Fatality rate (%) : The proportion of died patients.

**Figure 1.**
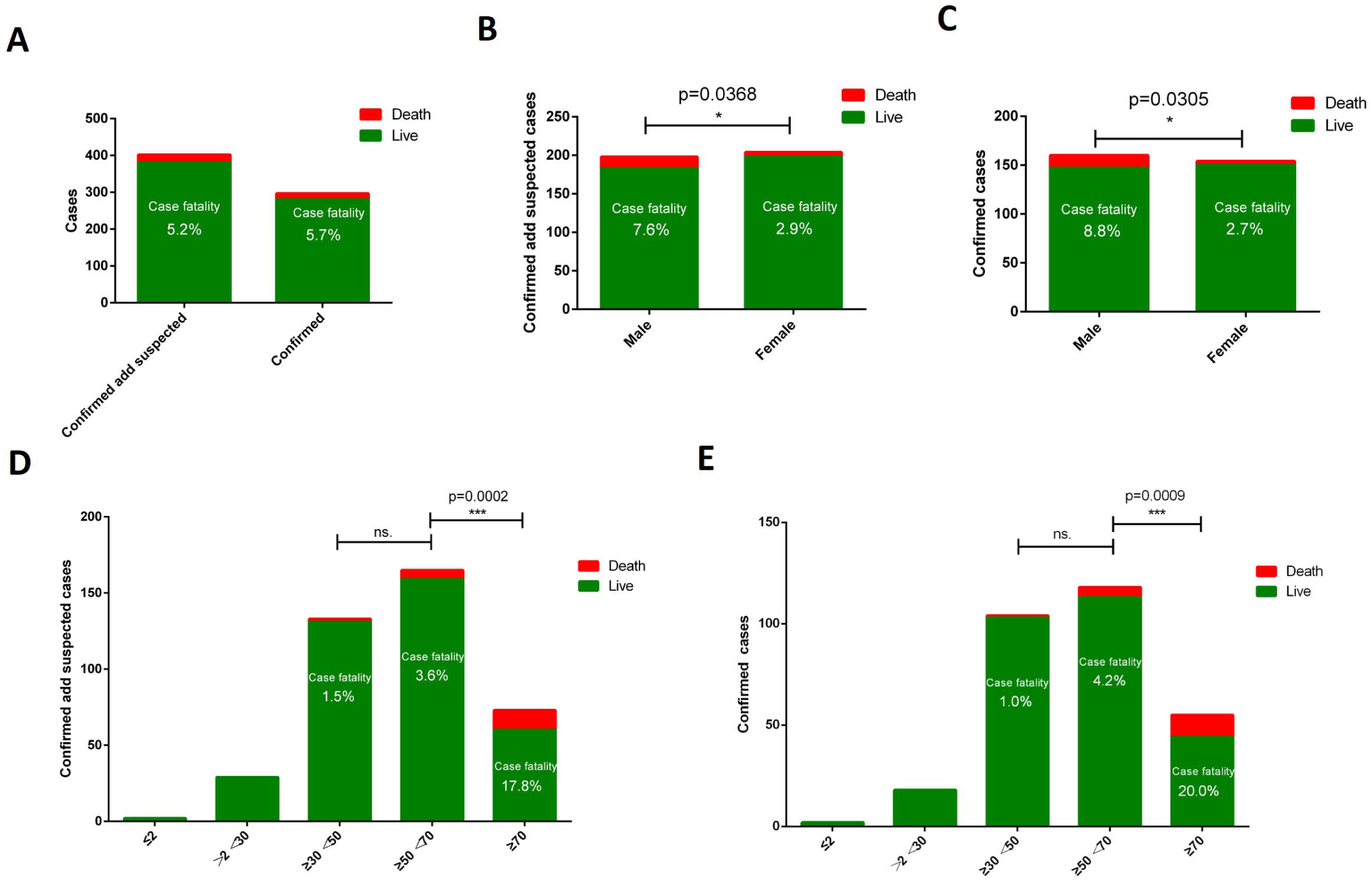
Case fatality analysis of SARS-CoV-2 patients. (A) Fatality of confirmed or suspected cases. (B) Male versus female patients among all suspected and confirmed patients. (C) Male versus female patients, confirmed cases only. (D) Fatality by age groups among suspected and confirmed patients. (E) Fatality by age group among confirmed patients. *P<0.05; ***P<0.001.

### Severity of illness among gender and age groups

As showed in **Table 2** and **Figure 2**, the overall severity rate (proportion of severe and critical severe cases, or SR) of confirmed and suspected cases was 35.1%, while the SR of confirmed cases was 27.3%. Male patients had a higher SR (38.9%) than female (31.4%) among suspected and confirmed patients, but the difference was not statistically significant (**Figure 2B**). In confirmed cases, male patients also had a higher SR than females without statistical significance (**Figure 2C**). The SR for patients under the age of 2 years, from 2 to 29, from 30 to 49, from 50 to 69 and over 70 was 0, 10.3%, 26.3%, 37.0% and 57.5% respectively, among all suspected and confirmed patients (Figure 2D). Which indicate that elder patients have higher severity rate than younger patients among all suspected and confirmed patients. The severity rate of patients under 2, from 2 to 29, from 30 to 49, from 50 to 69, 70 and over was 0, 11.1%, 17.3%, 28.8% and 52.7% respectively among all confirmed patients (**Figure 2E**). These results indicated that elder patients had higher severity rate than younger patients among all confirmed patients.

**Table 2.**
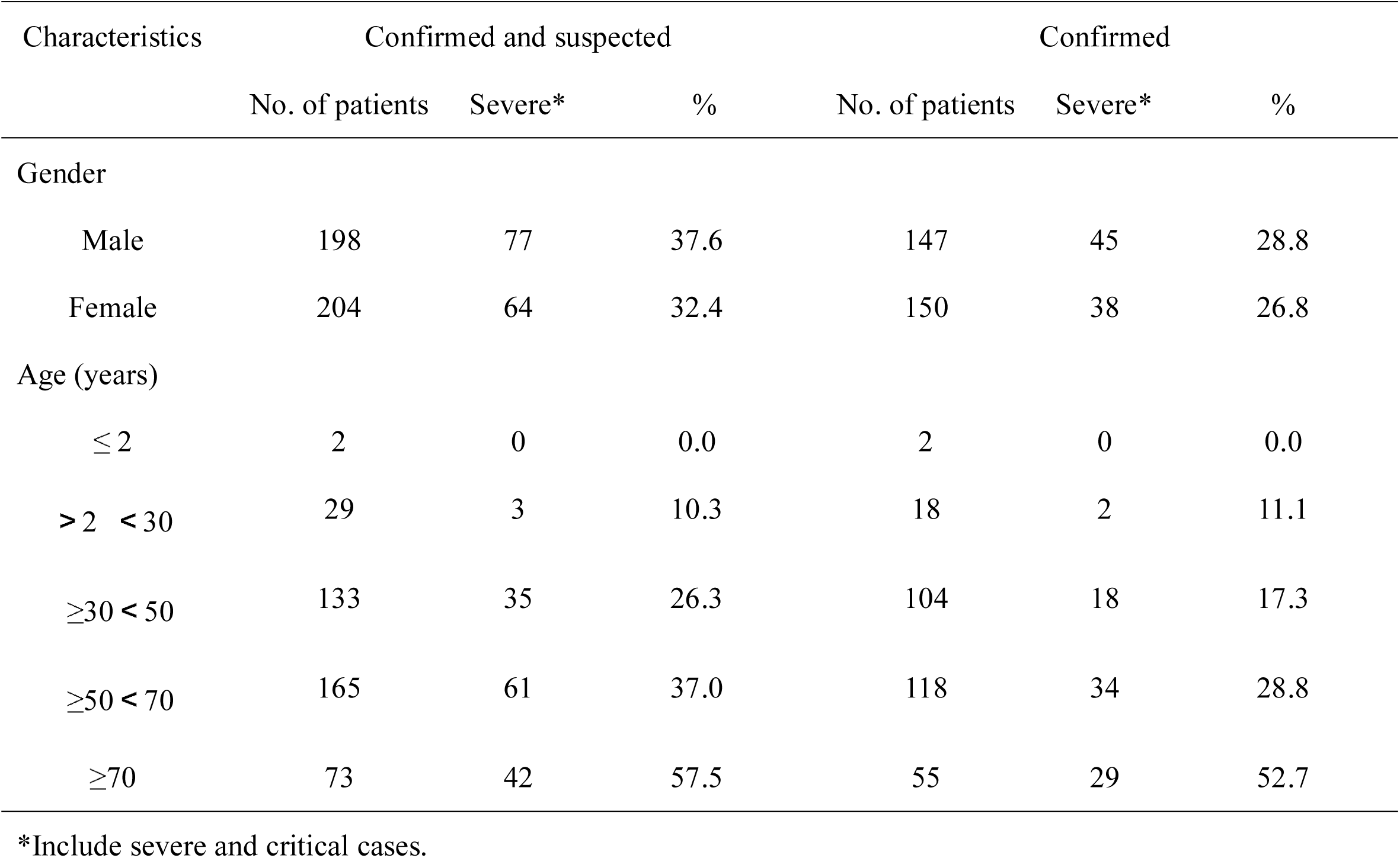
Portion of severe and critical cases by gender and age among patients with suspected and confirmed diagnosis of SARS-COV-2 infection.

**Figure 2.**
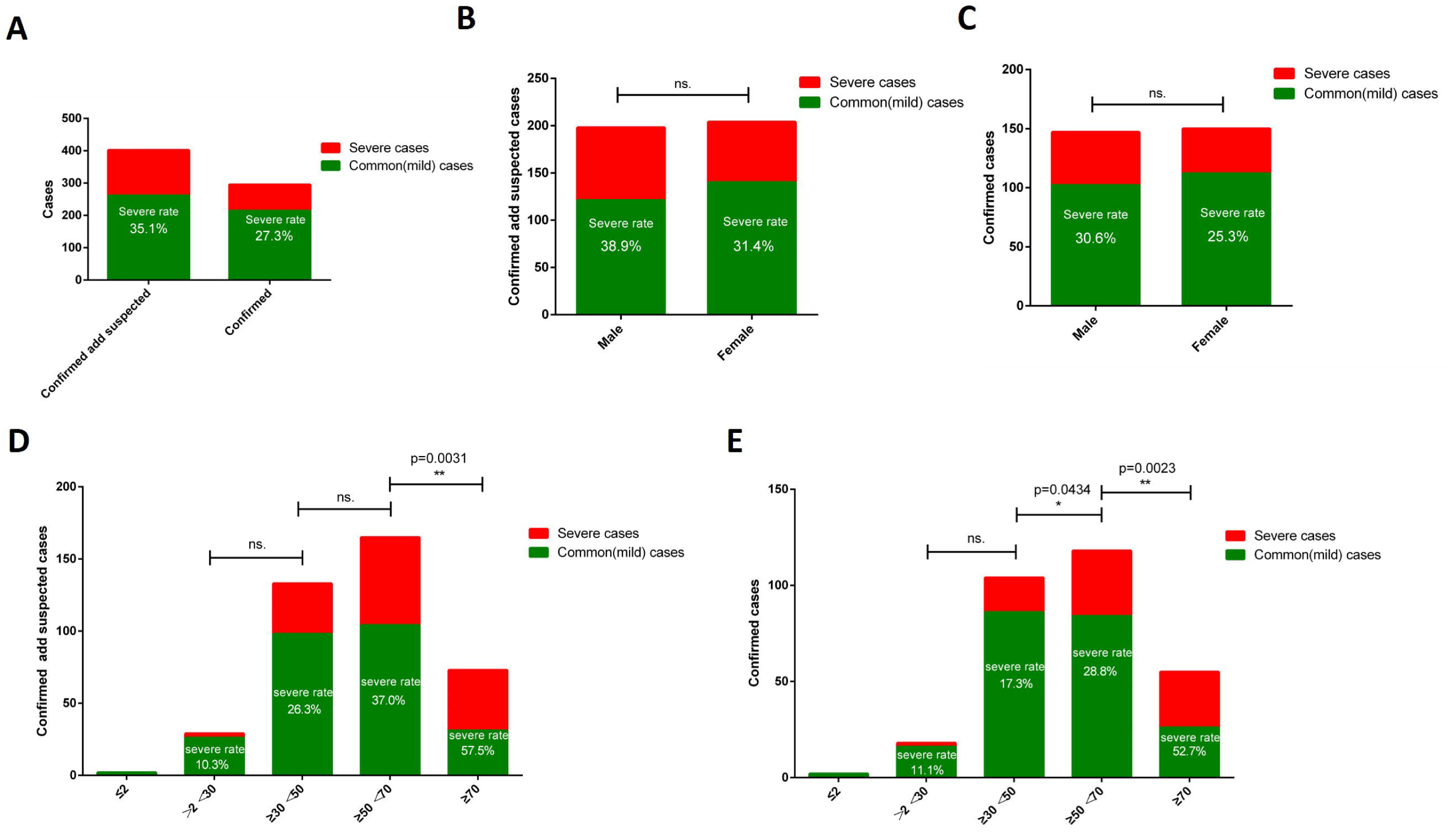
Clinical severity classification among SARS-CoV-2 patients by gender and age groups. (A) Overall severity classification in all cases versus confirmed cases. (B) Severity classification in male versus female patients among all patients. (C) Severity classification in male versus female patients among confirmed patients. (D) Severity classification by age groups among all patients. (E) Severity classification by age among confirmed patients. ns. no significance; *P<0.05, **P<0.01.

## Discussion

From late December, 2019 to February, 2020, the number of COVID-2019 patients is increasing in an astonishing speed. The symptoms of this disease include but are not limited to fever, cough, myalgia, diarrhea, dyspnea[1, 6]. Pathologically the lungs exhibit marked proteinaceous exudation and macrophages in alveolar spaces, as well as fibrin plugs in early phase [8], and hyaline membrane formation, reactive hyperplasia desquamation of alveolar epithelium [8, 9]. In addition to pneumonia, patients may also suffer injuries in the heart, liver and kidneys. Up till February 2, the mortality of this disease in Wuhan is 5.5%, and in Hubei is 3.2%, while in the region outside of Hubei is 0.1% (China National Health Commission official website).

This study included COVID-2019 patients from Zhongnan Hospital, one of the largest tertiary teaching hospitals in Wuhan, Hubei province. From late December, 2019 to early January, 2020, before large scale isolation measures were implemented, many departments in the hospital experienced cross-infection among patients and to the medical staff, due to unrecognized infectious patients and lack of proper protection. Many patients in incubation period came to the hospital for illnesses other than respiratory symptoms or fever[1]. After being designated as the specialized hospital, special isolation wards were built in each department[7]. The period from which we collected the clinical data was before the prevention was in full implementation. Indeed, the overall mortality and the rate of severe cases in the study are higher than the national figures. There may be several causes for the higher mortality in Wuhan compared to other cities. First, the rapid transmission of the virus and increase of patient volume quickly overwhelmed medical resources in Wuhan, leaving many patients to receive care later in the clinical course. Secondly, there were a large number of patients who had mild or transient symptoms who never received diagnosis nor nucleic acid tests, and were not counted as COVID-19 patients, making the denominator of mortality smaller than real.

We analyzed the mortality and proportion of the severe cases in gender and different age groups. Our cohort of 402 cases included patients of all ages, the youngest were a 1-month old girl and 6-month old boy, the oldest were a 94-year-old woman and a 96-year-old man. Most patients were between 30 and 80 years (87%) old, the median and average ages were 54 and 53 years, respectively. The highlights of findings are: (1) most deaths occurred in patients 70 years of age or older; (2) male patients had a mortality significantly higher than females (3 times); (3) no death occurred in patients 30 years of age or younger. It was shown that the higher mortality coincided with higher proportion in older age group having severe or critical illness. Patients with ages over 50 years were more susceptible to develop severe illness, particularly those in their 7^th^ decades. This is likely related to the fact that the majority of them had preexisting systemic illness. This finding is similar to that of influenza and SARS.

The difference in mortality between male and female genders is unknown. There appears that the severity distribution is equal between male and female groups. However, further analysis showed that mortality in severely ill male patients is higher than severely ill female patients. Some investigators reported that some chronic diseases are more common in males, such as hypertension[10], atherosclerosis, and chronic heart diseases[11]. But prevalence of these chronic diseases seems to be related to estrogen and equalize when women undergo menopause. Another possibility is that, expression of ACE2, the major receptor for the SARS-Cov-2, is higher expressed in males than in females[12]. But the study only included 8 individuals, with only 2 males (one Asian). Therefore, it is still a puzzle what roles ACE2 played in the pathogenesis of this catastrophic viral disease. Whether there is difference in viral load in males and in females, or whether more severe organ injury occurred in males, is still unknown. All these possibilities need further pathological and pathogenesis study. In addition, in a mouse models, it was found that males were more susceptible for SARS-CoV infection, although the result turned out that estrogen may have played a role[13].

Of note, we presented data in two different groups, one including the confirmed and suspected, and the other just the confirmed cases. As we know, during the outbreak case definitions had been changed, and application and interpretation of nucleic acid tests were not uniform for some period of time [14]. We believe it is best to see both the overall (including both confirmed and suspected) and confirmed group. Other than the individual figures in the results, the final conclusion remains the same in terms of comparisons between genders, and among different age groups. We strongly believe that all the suspected cases represent real COVID-19 patients, as they met the criteria for clinical diagnosis. Furthermore, as described in Methods, the interpretation of nucleic acid test subsequently removed the suspect category.

In conclusion, analysis of a cohort of 402 COVID-19 patients from a single center revealed an overall mortality of 5.2% and 17.8% mortality in patients 70 years of age or older. Male patients had a mortality 3 times that of female. No death occurred to patients age younger than 30. Causes for difference between male and female are currently unknown.

## Data Availability

All data will be available after publication

## References

1. Wang D, Hu B, Hu C, et al. Clinical Characteristics of 138 Hospitalized Patients With 2019 Novel Coronavirus-Infected Pneumonia in Wuhan, China. Jama 2020.

2. Zhu N, Zhang D, Wang W, et al. A Novel Coronavirus from Patients with Pneumonia in China, 2019. The New England journal of medicine 2020.

3. Chan JF-W, Kok K-H, Zhu Z, et al. Genomic characterization of the 2019 novel human-pathogenic coronavirus isolated from a patient with atypical pneumonia after visiting Wuhan. Emerging Microbes & Infections 2020; 9(1): 221–36.

4. Chan JF, Yuan S, Kok KH, et al. A familial cluster of pneumonia associated with the 2019 novel coronavirus indicating person-to-person transmission: a study of a family cluster. Lancet 2020; 395(10223): 514–23.

5. Li J, Ye, Guangming, Chen, liangjun, Wang, jiajun, Li, Yirong. Analysis of false-negative results for 2019 novel coronavirus nucleic acid test and related countermeasures. 2020; 43(00): E006–E.

6. Huang C, Wang Y, Li X, et al. Clinical features of patients infected with 2019 novel coronavirus in Wuhan, China. Lancet 2020.

7. Guan W-j, Ni Z-y, Hu Y, et al. Clinical characteristics of 2019 novel coronavirus infection in China. medRxiv 2020: 2020.02.06.20020974.

8. Tian S-f, Hu W, Niu L, et al. Pulmonary Pathology of Early Phase SARS-COV-2 Pneumonia. Preprints 2020, 2020020220 doi: 10.20944/preprints202002.0220.v1.

9. Xu Z, Shi L, Wang Y, et al. Pathological findings of COVID-19 associated with acute respiratory distress syndrome. The Lancet Respiratory Medicine 2020.

10. Raghvendra K. Dubey SO, Bruno Imthurn, Edwin K. Jackson. Sex hormones and hypertension. cardiovascular research 2002; 53(3): 688–708.

11. Denton KMMCKM. Sex-specific differences in hypertension and associated cardiovascular disease. nature reviews nephrology 2018; 14(3): 185–201.

12. Zhao Y, Zhao Z, Wang Y, Zhou Y, Ma Y, Zuo W. Single-cell RNA expression profiling of ACE2, the putative receptor of Wuhan 2019-nCov. 2020.

13. Channappanavar R, Fett C, Mack M, Ten Eyck PP, Meyerholz DK, Perlman S. Sex-Based Differences in Susceptibility to Severe Acute Respiratory Syndrome Coronavirus Infection. The Journal of Immunology 2017; 198(10): 4046–53.

14. Xiao S-Y, Wu Y, Liu H. Evolving status of the 2019 novel coronavirus infection: Proposal of conventional serologic assays for disease diagnosis and infection monitoring. J Med Virol 2020.

